# Biosimilarity of GBPD002 compared with Eprex^®^ through clinical evaluation in human

**DOI:** 10.1101/2023.01.29.23285155

**Authors:** Kakon Nag, Mohammad Mohiuddin, Mamun Al Mahtab, Sitesh Chandra Bachar, Abdur Rahim, Helal Uddin, Samir Kumar, Maksudur Rahman Khan, Enamul Haq Sarker, Mashfiqur Rahman Chowdhury, Rony Roy, Sourav Chakraborty, Bipul Kumar Biswas, Emrul Hasan Bappi, Ratan Roy, Uttam Barman, Naznin Sultana

**Affiliations:** Globe Biotech Limited, 3/Ka (New) Tejgaon I/A, Dhaka 1208, Bangladesh; R&D Management Solution Inc., Hamilton, Ontario L9C2V8, Canada; Department of Hepatology, BSMMU, Dhaka, Bangladesh; Faculty of Pharmacy, University of Dhaka, Dhaka, Bangladesh; Clinical Research Organization Ltd. (CRO Ltd.), Dhaka, Bangladesh

**Keywords:** Erythropoietin, biosimilarity, pharmacokinetics, pharmacodynamics, toxicity

## Abstract

**Background:** The biosimilarity for erythropoietin (EPO) functionality of GBPD002 (test candidate) and Eprex^®^ (comparator) has been evaluated by comparing the pharmacokinetic (PK) and pharmacodynamic (PD) properties following subcutaneous injection.

**Methods:** This was a randomized, double-blinded, two-sequence, crossover clinical trial. Subjects were randomly assigned and received a dose (4,000 IU) of either the test or comparator EPO, and received the alternative formulations after 4-weeks of washout period.

**Results:** The PK parameters, *viz*., maximum observed concentration (C_max_) and area under the curve extrapolated to infinity (AUC_0-inf_), were calculated with the serum EPO concentrations from blood samples and were found comparable for both formulations. The geometric mean ratios (at 90% CI) of the C_max_ and AUC_inf_ were 0.89 and 1.16, respectively, which were within the regulatory range of 0.80 – 1.25. The time-matched serum EPO concentrations and PD markers (reticulocyte, hematocrit, hemoglobin, and red blood cell) denoted a counterclockwise hysteresis, suggesting a time delay between the observed concentration and the response. ANOVA-derived *P-*values (>0.05) for the effectors clearly revealed the similarity between effects on PD markers for the test and comparator drugs. Both formulations were found tolerated well, and anti-drug antibodies were not observed.

**Conclusions:** Thus, the two formulations are projected to be used interchangeably in clinical settings.

## Background

Erythropoietin (EPO) is a glycoprotein hormone that plays a key role in the formation of red blood cells (RBCs) [1]. EPO is primarily synthesized in the peritubular cells of the kidney and released into the systemic circulation in adult individuals [2]. Circulating EPO binds to the EPO receptor on bone marrow erythroid progenitors, triggering multiple signaling pathways that support differentiation into mature RBCs [3]. A reduction in EPO production is the primary cause of anemia in people with chronic renal failure [4]. Human recombinant epoetin (rHuEPO) or erythropoiesis-stimulating agents (ESA) have been demonstrated to stimulate erythropoiesis in anemic patients with chronic renal failure, including those who need and don’t need dialysis [5 – 7]. Hereafter, EPO and rHuEPO have been used in this article interchangeably. ESAs are used to treat chemotherapy-induced anemia in cancer patients and to reduce the requirement for allogenic blood transfusions in patients with mild anemia who are undergoing surgery [8 – 10]. Furthermore, EPO is recommended for patients who are at high risk for perioperative transfusions due to considerable blood loss.

Alpha epoetins are the most commonly used type of rHuEPO among the other forms. Despite its tremendous importance in the clinical field, the price of EPO remains significantly high, and limiting its availability to the mass people – particularly, to the underdeveloped and developing countries. Eprex^®^, the pioneer product of alpha epoetins, is a regular medicine with proven efficacy and tolerability [11]. Eprex^®^ was manufactured by Johnson & Johnson and it was the first EPO formulation to receive regulatory approval in Europe in 1988. In the early 1990s, physicians outside of the United States adopted the subcutaneous route of administration of EPO for hemodialysis patients due to the socio-economic benefit for the patients [11]. Human serum albumin (HSA), the stabilizer in Eprex^®^ formulation, was changed to a synthetic compound, polysorbate 80, due to the concerns that albumin might transmit Creutzfeldt-Jakob disease [11]. Subsequently, HSA-free Eprex^®^ has been available in market.

GBPD002 is a biosimilar of Eprex^®^, which is developed by Globe Biotech Limited and synthesized in genetically engineered Chinese hamster ovary (CHO) cells. Upstream and downstream process development and validation were done for large-scale production [12]. Step-by-step analytical results confirmed the biosimilarity of GBP002 with Eprex^®^ regarding molecular characterization [12]. Single and repeat-dose toxicity was performed in Wister rats to analyze the toxicity of GBPD002 with Eprex^®^ and were found safe for administration. Several PK/PD studies were performed in animal models and results were found to be similar for GBPD002 and Eprex^®^ [12].

Here, the aim of this study is to analyze the bioequivalence of GBPD002 and Eprex^®^. The purpose of this study is to compare the pharmacokinetic (PK), pharmacodynamics (PD), and safety of rHuEPO, GBPD002, developed by Globe Biotech Limited, and the reference product Eprex^®^ manufactured by Janssen Cilag Ltd., UK in healthy adult volunteers.

## Materials and methods

### Study design

A randomized, double-blind, single-dose, and two-sequence crossover trial in healthy volunteers was designed. The protocol for the study has got ethical clearance from the institutional review board (IRB; The Ethics Committee of Farabi General Hospital Ltd.) and was approved by the Directorate General of Drug Administration (DGDA) of Bangladesh [13]. The Clinical Research Organization (CRO Ltd.) conducted this investigation at Farabi General Hospital, Dhanmondi R/A, Dhaka 1209, in compliance with the principles of the Declaration of Helsinki and the International Conference on Harmonization’s Guideline for Good Clinical Practice. The informed consent form (ICF), along with the protocol and methodology, was also approved by IRB and DGDA. Volunteers were given thorough information about the risks and benefits of the study and they signed the ICF to affirm their willingness to voluntarily participate in the study. The trial protocol has been submitted and registered with the clinicaltrial.gov of the National Library of Medicine of the USA [14].

Briefly, the study was open to healthy male participants aged 19 – 45 years old who weighed 55.0 – 90.0 kg and had a body mass index (BMI) of 18.0 – 27.0 kg/m^2^. Subjects were eliminated if they had at least one of the following clinical laboratory test results: hemoglobin level <12 g/dL or >17 g/dL, vitamin B12 level <200 pg/mL, ferritin level <21.8 ng/mL, transferrin level <190 mg/dL and any anomalous range for the reticulocyte (RET) count, erythrocytes, platelets or serum potassium levels. The study design is supported by a previously accomplished similar study [15]. The total number of subjects was 42, assuming a 20% dropout rate. HIV, HBsAg, and HCV (Hepatitis C Virus) positive individuals were excluded from the study.

The study was conducted after the emergence of COVID-19, and therefore, proper precautions have been taken to protect the volunteers against exposure from COVID-19 during the trial. For example, during the primary selection, admission into the trial site, interaction during sampling and monitoring (including periods for waiting, washroom usage, food and drink intake, entry and exiting, housekeeping, etc.) were strictly controlled. The usage of approved masks and hand sanitizers was mandatory during the stay at the trial facility. All relevant people including investigators, medical doctors, nurses, analysts, support staffs, etc. who were engaged in the trial and with an opportunity to access the trial site were controlled; all of them followed the same rigorous access and mobility protocol. The volunteers were housed in isolated cabins where maximum of 2 persons were allocated in a single room in separate beds located at a minimum of 6 feet distance to reduce the frequency of interaction between subjects and interacting people. Regular COVID-19 tests were included for relevant sampling points, and the subjects with positive test results were immediately isolated and subjected to proper medical care.

The recruited volunteers were randomly assigned to one of the two sequences (Sequence/Group A and Sequence/Group B) and received a single subcutaneous injection of 4,000 IU of either the comparator/reference drug (Eprex^®^) or the test drug (GBPD002) in the abdomen area from a single-use prefilled syringe (PFS; Schott syriQ BioPure^®^), SCHOTT Schweiz AG, Switzerland). The allocated sequences with a 28-day washout period were as follows: Group A, administered the comparator drug in period 1 followed by the test drug in period 2; Group B, given the test drug in period 1 followed by the comparator drug in period 2.

Blood samples were taken for the PK evaluation at predose and at 1, 3, 6, 8, 10, 14, 24, 48, 72, 96, 120, and 144 h postdose. For the PD evaluation, the reticulocyte count (RET, %), hematocrit (HCT, %), haemoglobin (HB) (g/L), and red blood cell (RBC) count (10^6^/mm^3^) were calculated at predose and at 72, 144, 216, and 312 h postdose.

To maintain iron supply, instead of iron supplements in the form of medicine, the subjects were given a standard portions of iron-rich food prepared with green leafy vegetables, animal organs like the liver and red meat, beans, pumpkins etc. Watermelon, dried dates and, dark chocolates were provided as snacks.

### Bioanalytical methods

A validated enzyme-linked immunosorbent assay (ELISA) technique was used to measure serum EPO quantities. *Quantikine*^*®*^ *IVD*^*®*^ *ELISA, a human EPO* immunoassay kit (R&D Systems Inc., Minneapolis, MN, USA) was used to determine the serum EPO concentrations. Exogenous EPO derived from rHuEPO and endogenous EPO were measured together in the same way. The procedure was validated following international guidelines. The calibration curve was constructed using seven distinct concentrations of calibration standard samples. Samples of low to high concentrations (2.5, 5, 20, 50, 100, and 200 mIU/mL) were prepared for quality control. Calibration curves of the test and comparator drugs showed linearity, (r^2^>0.99 for both test and comparator) within the concentration range of 2.5 – 200 mIU/mL. The hematologic parameters for PD assessment (RET, HCT, HB, and RBC count) were analyzed in a diagnostic center (Lab Science Diagnostic), which is accredited by the Directorate General of Health Sciences (DGHS) of Bangladesh.

### PK and PD analyses

The PK parameters were measured by a noncompartmental method using Phoenix^®^ WinNonlin^®^ (Version 8.3; Certara, L.P., Princeton, NJ, USA). Raw data were used to determine the maximum observed serum EPO concentration (C_max_) and the time of C_max_ (T_max_). The last observed area under the curve (AUC_last_) was determined by the linear trapezoidal method up to T_max_ and by the log trapezoidal method after T_max_. The area under the curve was extrapolated to infinity (AUC_inf_) with the following formula: AUC_inf_ = AUC_last_ + C_last_/λ_z_, where C_last_ refers to the last observed serum EPO concentration and λ_z_ refers to the estimated terminal elimination rate constant. The terminal half-life (t_1/2_) was calculated by dividing natural-log 2 by λ_z_. The total clearance (CL/F) was determined with the following formula: CL/F = dose/AUC_last_, where F means the bioavailability. The mean residence time (MRT_last_) was calculated by dividing the area under the first moment curve by the AUC_last_ [13]. Goodcalculator™ and socscistatistics™ were used respectively to determine 90% CI and *P* value.

As PD indicators, the time courses of RET count, HB, HCT, and RBC count were studied and compared between the test and comparator drugs. The linear trapezoidal approach was used to determine the maximum effect change (Emax) and the area under the baseline-adjusted effect curve (AUEC) for the RET count, HB, HCT, and RBC count using baseline-adjusted values. We analyzed inter-and intra-volunteer data trends for all parameters and outlier data were excluded from the final analysis to minimize errors in data prediction [16]. The time-matched PK/PD data (serum EPO concentration and each PD marker) were also plotted on a scatter plot to investigate the PK/PD time delay.

### Safety and tolerability analysis

Safety and tolerability profiles of the drugs were evaluated in participants who had at least one dose of the study drug. The results of vital sign evaluations, electrocardiograms, and clinical laboratory testing were used to determine the safety and tolerability of the drug. Local reactivity for drug administration was assessed 1, 24, and 48 hours after injection. Anti-drug antibody (ADA) production was measured at the predose of each period and at the post-study visit to determine the immunogenicity of the study drugs.

### Statistical analysis

The key PK parameters C_max_ and AUC_inf_ were used in the PK comparison. A linear mixed-effect analysis of variance was used to calculate the log-transformed C_max_ and AUC_inf_, with a fixed effect for the formulation, period, and sequence and a random effect for the subject nested for the sequence. For each PK parameter, the geometric mean ratio (GMR) of the test to the comparator was determined, along with its 90% CI. If the 90% CI for each PK parameter was within the range of 0.80 – 1.25, the test drug was determined to have PK equivalence with the comparator drug. The key PD parameters, E_max_ and AUEC of the RET count, were included in the PD comparison. The mean difference between the test and the comparator drug was calculated using the linear mixed-effect analysis of variance, along with its 90% CI and P-value. Statistical significance was defined as a *P*-value of less than 0.05; the *P-*value(s) for PD marker(s) for the EPO formulations greater than 0.05 were considered similar and non-significant.

## Results

### Demographics

Total of 83 persons were screened to include 42 volunteers (50.6% were eligible). The number of the study subject has been aligned with several other studies to achieve significant data collection and confident decision-making [16 – 20].

It has been shown that comparatively a lower dose provides more stable PK/PD results than the higher dose [21, 22]. Several studies have administered 4000 IU dose for bioequivalence studies as an effective and comparably low dose, [15, 23], and accordingly, we have used 4000 IU/subject as the experimental dose. One volunteer from group A (started with comparator drug) and two volunteers from group B (started with test drug) dropped out from the study after the first phase (28 days) and before receiving the second dose due to being positive for COVID-19. The mean ±SD (min–max) for age, height, weight, and BMI for the volunteers were measured and shown in Table 1. The demographic and other baseline variables were found not significantly different among the two groups.

**Table 1.** Demographic characteristics of the study groups.

| Demographic Characteristics | Sequence/Group A (n=21) | Sequence/Group B (n=21) |
| --- | --- | --- |
| Mean Age (years) | 27.67±6.03 | 31.57±6.90 |
| Mean Height (inch) | 64.4±0.21 | 64.1±0.17 |
| Mean Weight (kg) | 59.94±6.50 | 64.88±6.00 |
| Mean BMI (Kg/m<sup>2</sup>) | 22.4±2.21 | 24.5±3.35 |
| SBP (mmHg) |  |  |
| *Baseline* | 115.5±5.41 | 122.78±7.45 |
| *Endpoint* | 115.6±6.96 | 123.13±10.24 |
| DBP (mmHg) |  |  |
| *Baseline* | 75.0±4.26 | 79.44±1.49 |
| *Endpoint* | 73.20±6.04 | 81.13±5.09 |

### PK results

After a single subcutaneous injection of both formulations, the amplitudes of serum EPO concentrations followed comparable time-dependent distributions. EPO demonstrated a delayed systemic absorption with a median T_max_ of 6 – 8 h in both formulations and displayed multiphasic characteristics in the elimination phase (Figure 1). The GMR (test/comparator) for both the C_max_ and AUC_inf_ fell within the pre-specified range of 0.80–1.25 (0.89 and 1.16, respectively), suggesting that the two epoetin alfa formulations have similar PK profiles. The remaining PK characteristics were similar between the two formulations as well and shown in Table 2.

**Table 2.**
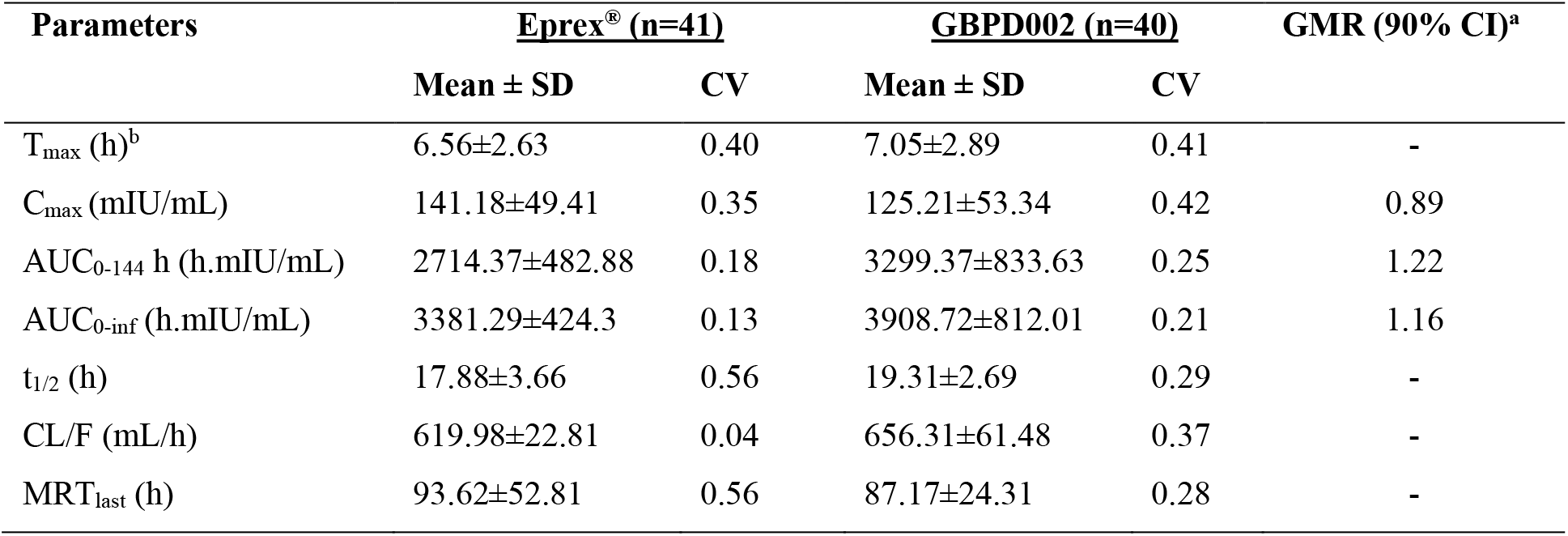
Pharmacokinetic parameters after a single subcutaneous administration of the test or the comparator drug products.

**Fig. 1.**
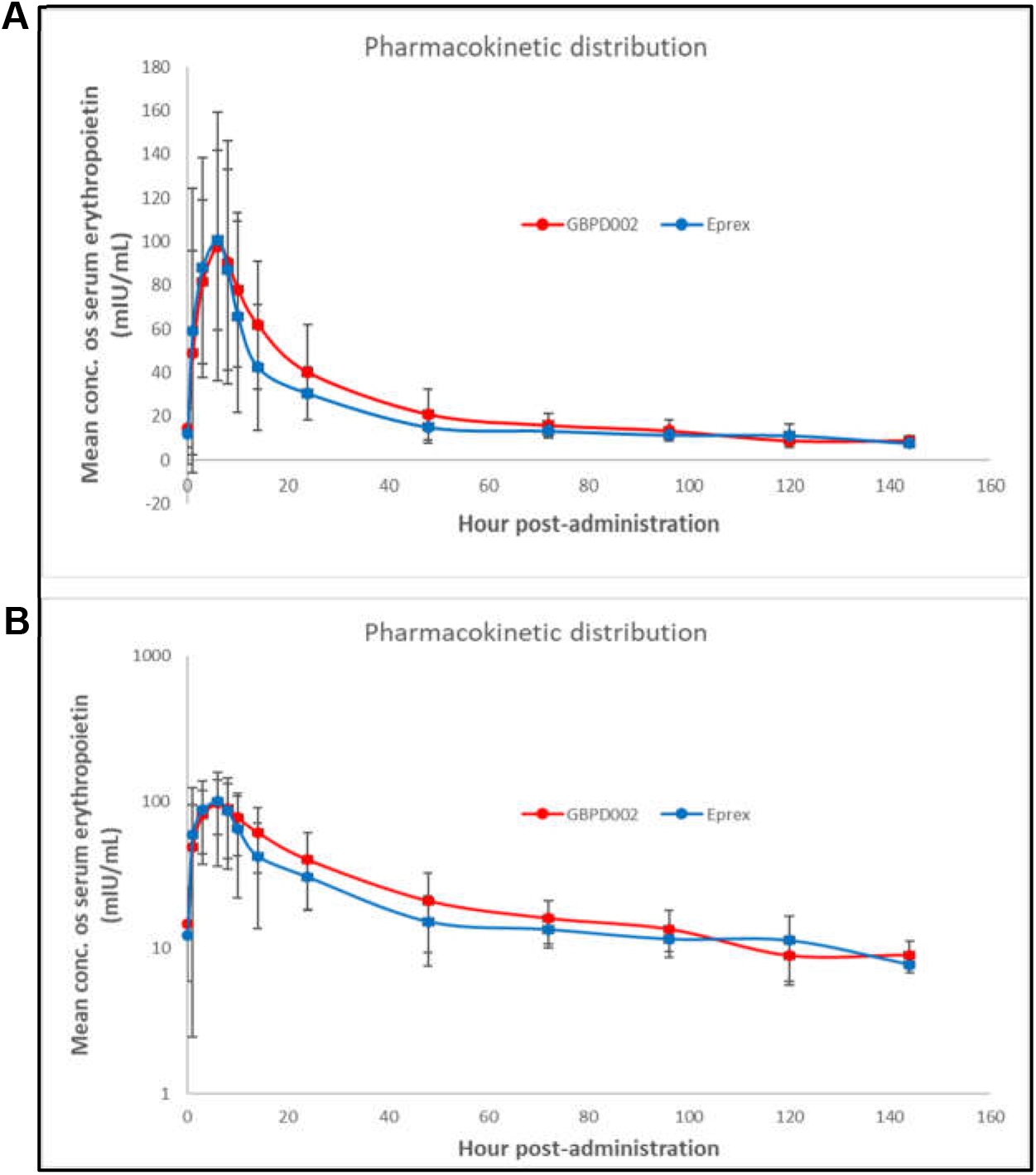
Time-dependent mean serum concentration profile of serum EPO after a single subcutaneous injection of the comparator (Eprex^®^, blue circle) or test (GBPD002, red circle) epoetin alfa. **A**) liner numerical scale, **B**) log scale.

### PD results

The mean RET counts progressively increased up to 72 hours following drug administration and declined until the last observation time (at 336 h). The test and comparator EPO had a similar rates of RET count change over the observed time span. The key PD parameters, *viz*., mean E_max_ and AUEC of the RET count, were comparable between the test and the comparator (*P*=0.604 and 0.976, respectively). Furthermore, count changes, E_max_, and AUEC were similar for HB, HCT, and RBC between the two formulations (Figure 2 and Table 3). The similarities in PD parameters between the comparator and the test drug were clearly evident from the non-significant *P*-values, which are above 0.05 for each PD parameter.

**Table 3.** Pharmacodynamic parameters after a single subcutaneous administration of the test and comparator drug products.

| Parameters | Eprex® (n=41) |  | GBPD002 (n=40) |  | GMR<br>(90% CI)<sup>a</sup> | P-value<br>(ANOVA) |
| --- | --- | --- | --- | --- | --- | --- |
|  | Mean ± SD | CV | Mean ± SD | CV |  |  |
| Reticulocyte |  |  |  |  |  |  |
| T<sub>max</sub> (h)<sup>b</sup> | 108.48 ±2.70 | - | 127.2±2.81 | - | - | - |
| Δ E<sub>max</sub> (%) | 91.94 ± 46.04 | 0.51 | 96.29 ± 40.88 | 0.42 | 1.047 | 0.604 |
| Δ AUC (h.%) | 642.29 ± 328.85 | 0.53 | 704.59 ± 115.0 | 0.16 | 1.097 | 0.976 |
| Hemoglobin |  |  |  |  |  |  |
| T<sub>max</sub> (h) | 118.56±3.61 | - | 86.4±1.20 | - | - | - |
| Δ E<sub>max</sub> (g/dL) | 5.45 ± 3.32 | 0.61 | 4.90 ± 1.73 | 0.34 | 0.901 | 0.747 |
| Δ AUC (h. g/dL) | 37.9 ± 14.50 | 0.38 | 37.86 ± 11.60 | 0.31 | 0.999 | 0.859 |
| Hematocrit |  |  |  |  |  |  |
| T<sub>max</sub> (h) | 122.4±3.62 | - | 123.36±3.84 | - | - | - |
| Δ E<sub>max</sub> (%) | 4.57 ± 3.71 | 0.48 | 4.55 ± 2.64 | 0.58 | 0.999 | 0.970 |
| Δ AUC (h.%) | 95.57 ± 64.38 | 0.67 | 96.46 ±57.08 | 0.59 | 1.009 | 0.572 |
| Red blood cell count |  |  |  |  |  |  |
| T<sub>max</sub> (h) | 126.24±3.45 | - | 76.32±3.38 | - | - | - |
| Δ E<sub>max</sub> (10<sup>6</sup>/mm<sup>3</sup>) | 6.89 ± 2.69 | 0.52 | 6.07 ± 3.42 | 0.56 | 0.890 | 0.604 |
| Δ AUC (h. 10<sup>6</sup>/mm<sup>3</sup>) | 43.67 ± 28.08 | 0.64 | 43.35 ± 30.54 | 0.70 | 0.993 | 0.976 |

**Table 4.**
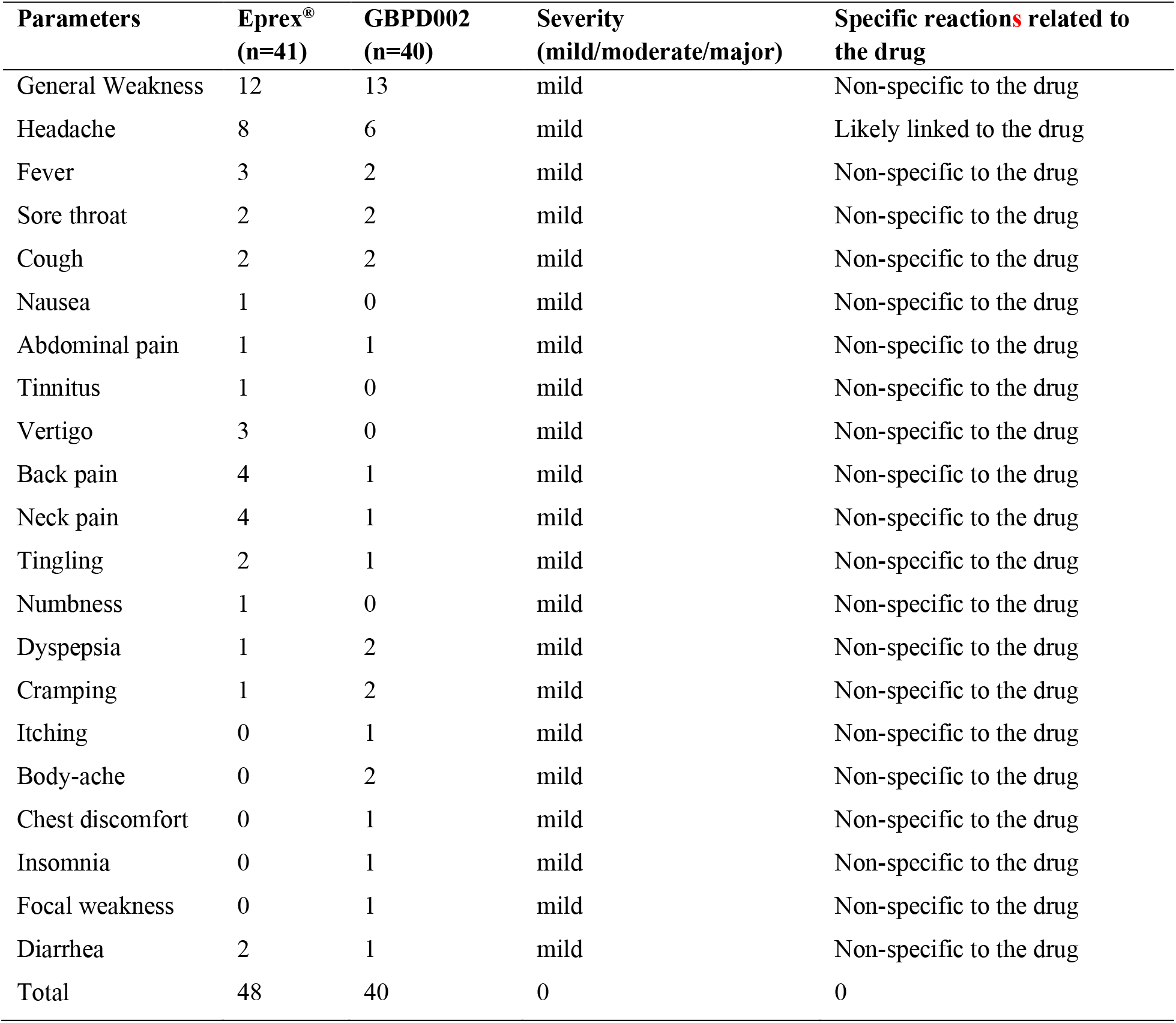
Reported adverse events during the course of the study.

**Fig. 2.**
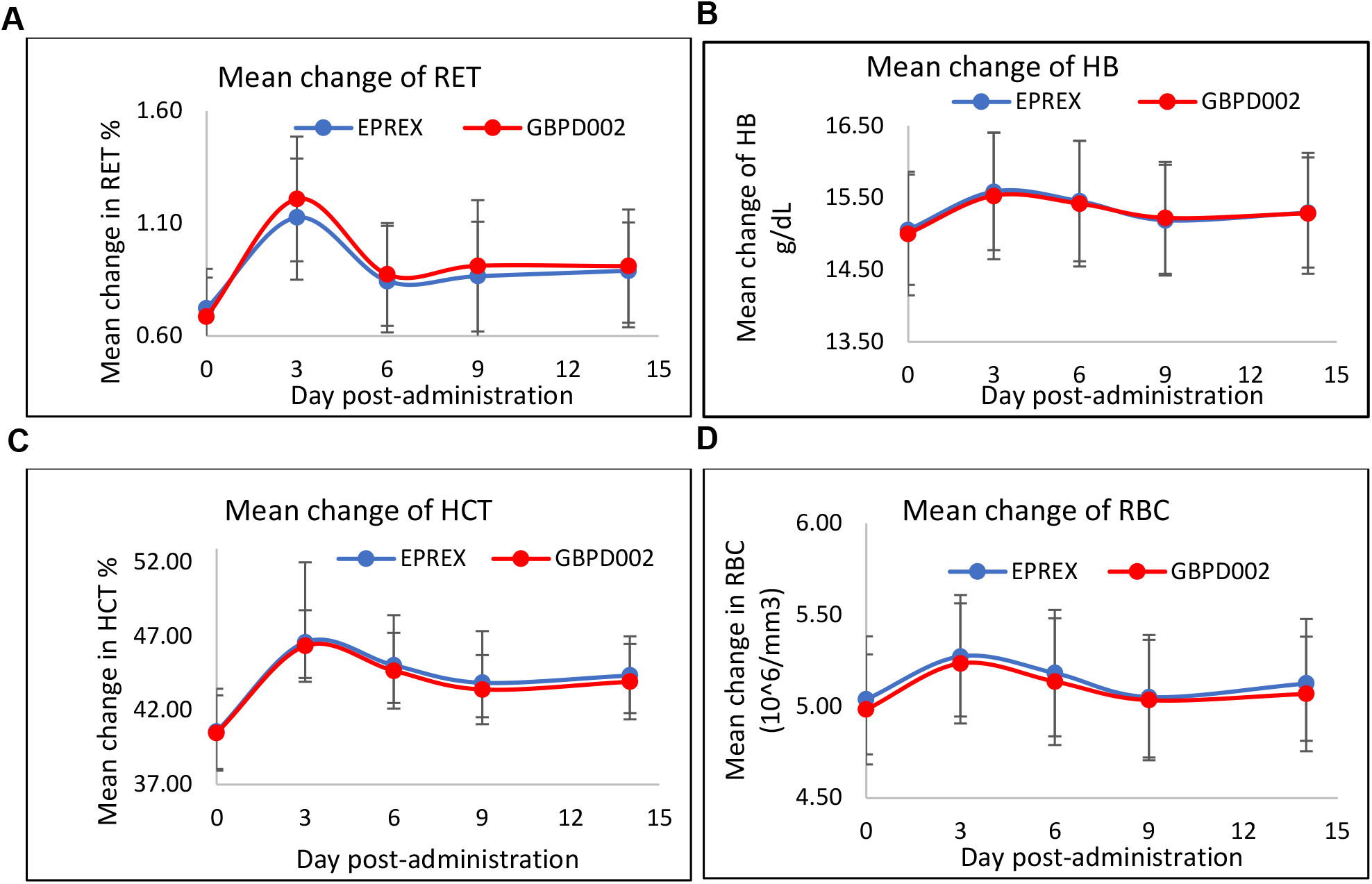
Mean change in hematologic parameter levels following a single subcutaneous administration of the test (blue circle) or the comparator (red circle). **A**) RET, **B**) HB, **C**) HCT and **D**) RBC.

### Safety and tolerability

After receiving a single subcutaneous injection of 4,000 IU EPO, 27 subjects reported 88 adverse events (AE). The AEs were addressed immediately by the medical team as per protocol and recorded. Between the two treatments, the number of patients with AEs and the number of AEs were comparable. All the treatment-related AEs were mild in severity and did not require any medication. General weakness and headache were the most commonly reported treatment-related AEs, which are already recognized to be common mild side effects of rHuEPO formulations [24]. Clinical laboratory results, ECG readings, vital signs, and physical tests all showed no clinically significant changes for receiving two doses (test and comparator) of EPO. No local reaction was observed on the injection site among the volunteers. No ADA reactivity was found in any samples from both of the treatment groups.

## Discussion

The PK and PD parameters of two EPO formulations were evaluated and compared in this study. It has been shown that the pharmacological response for PD markers (RET, HB, and RBC) were sex independent [25]. Therefore, in our study, we have included only male volunteers. Yoon S *et al*. have recently performed the PK and PD study between a candidate EPO formulation with Eprex^®^ in healthy male volunteers [15]. They have found comparable responses between the test and comparator for PK and PD parameters, and extrapolated their study findings sex-independently. In our study, we found that the PK and PD profiles and values of the test EPO (GBPD002) were similar to those of the comparator EPO (Eprex^®^).

EPO is biologically removed from systemic circulation by attaching to its cognate receptor in the bone marrow [27]. The t_1/2_ data in our study is in close proximity to other comparable studies [17 – 23], suggesting that the variables of the study were in coordination with other studies performed in different study sites and subjects. Previous studies reported that the PK of epoetin alfa may be influenced by the dynamic binding characteristics showing nonlinear disposition profiles [15]. In this study, a typical nonlinear elimination tendency was observed accordingly for both formulations. Other EPO receptor-binding drugs, such as epoetin beta and darbepoetin alfa, have also shown similar elimination patterns [28, 29].

The RET count was shown to be highly predictive of erythropoiesis efficacy [30]. The dose-response association for the RET count with rHuEPO has been well established [31]. Further, the RET count has been recommended as the key PD marker in European and other recommendations for single-dose SC administration trials for EPO [32]. Therefore, RET count was determined as the key PD marker in this investigation. Reticulocyte populations increased as the bone marrow resident hematopoietic cells responds to EPO. A RET number of fewer than 10,000/μl is considered to represent no or minimal regenerative response, 10,000–60,000/μl is a poor regenerative response, 60,000–200,000/μl is moderate response and 200,000–500,000/μl is maximal regenerative response [33]. We have observed that the experimental formulation of EPO induced RET population significantly by day 3 and comparably with the reference product. RET population came down to the basal level on day 6, which is in accordance with the fact that the circulating RET population transformed to RBC within 2/3 days [34, 35]. This notion was supported by the findings that the hematocrit (HCT) population went up on EPO administration on day 3, and the level was sustained after a slight dip. This data clearly suggested the classical effectiveness of both EPO preparation used in the study for RBC generation through the RET-induction pathway.

Though minor and insignificant but a trend of very little higher response was observed for RET data for GBPD002 than the Eprex^®^. This observation can be assigned to the fact that the EPO function is dependent on resident time in the system rather than the higher level of concentration at a certain time point [36]. We also observed that the GBPD002 level remains a little higher in the system over the Eprex^®^, which is supportive to the notion.

HB has been considered as another PD marker for EPO in several clinical studies. In our study, HB levels were found increased by 0.5g/dl on day 3 after the SC administration of EPO and then declined to a normal level. Our result is in accordance with the findings of other studies where a similar level of raise in HB level was observed for SC-routed EPO administration [19, 22, 37, 38]. The RBC reading for our study also has shown similar results to the findings of Krzyzanski *et al*., and Sorgel *et al*., where the RBC level rises at a similar level on day 3 and then fell down to the basal level within 5/6 days [38, 39]. Yan *et al*., did not include RBC count in their study [40], therefore, a comparison of RBC response with their study was not possible. However, HB levels in all these studies and RBC levels in Krzyzanski *et al*., and Sorgel *et al*.*’s*, studies went up steadily after the administration of follow-on repeat doses. Considering very resembling trends for these PD markers for single-dose in duplicate segments, it can be expected that the responses for follow-on multiple doses for GBPD002 would be responding alike. Collectively, these results shown here demonstrated similar responses for PD markers for experimental EPO preparations, *viz*., GBPD002 and Eprex^®^.

Counterclockwise hysteresis was seen for both treatments when serum EPO concentrations and PD marker levels were time-matched. A counterclockwise hysteresis refers to a time delay between the measured concentration and the PD reaction. The duration of exogenous EPO migrating from systemic circulation to its binding site in the bone marrow, as well as the delayed detection of the response, are possible sources of this indirect association [41, 42]. More specifically, the maturation of normoblasts into RET takes 5 – 7 days, and EPO plays a key part in this process [43].

No ADA was developed during the study period for either formulation. The main safety concern for EPO administration is the risk for the development of antibodies, which consequently develops epoetin-associated pure red cell aplasia (PRCA). EPO-associated PRCA was first reported in 1998 [44], and is characterized by severe anemia, low RET count, erythroblast absence, EPO nonresponse, and neutralizing antibodies [45 – 47]. Later the cause for antibody generation was attributed to leachates from the rubber stoppers and corrected by the application of Teflon coating to the rubber of the stopper [48]. The incidence of EPO-associated PRCA has significantly reduced thereafter [49,50]. In our study, we could not detect EPO-specific antibodies in any of the samples. Previous studies have also reported that ADA development is not common after subcutaneous injection of epoetin alfa [15], which has been suggested that non-specific immunoreactogenicity is unlikely to happen for the EPO formulation under test. We have used a special polymer-coated leachate-free rubber stopper for dose presentation in single-use PFS. Therefore, long-term risk for the generation of EPO-mediated PRCA from GBPD002 preparation is highly unlikely.

In conclusion, the PK and PD profiles of the test (GBPD002) epoetin alfa were identical to those of the comparator (Eprex^®^). The two medicines had similar levels of tolerability, including local toxicity and immunoreactogenicity profiles. Between the two products, the hysteretic connections between serum EPO levels and erythropoietic responses were similar. Because the two epoetin alfa medications have comparable pharmacokinetic, pharmacodynamics, and safety profiles therefore they can be considered biosimilar in clinical settings, and likely be administered interchangeably.

## Data Availability

All data produced in the present work are contained in the manuscript

## Abbreviations

CV: coefficient of variation
SD: standard variation
CI: confidence interval
GMR: geometric mean ratio ^a^GMR: (90% CI) of the test to the comparator epoetin alfa
^b^ T_max_ (h): Mean (lowest-highest)
AUC_0–144_ h: area under the curve from time zero to the time of the last observation
AUC_0-inf_: area under the curve extrapolated to infinity
C_max_: maximum observed serum EPO concentration
CL/F: total clearance
MRT_last_: mean residence time
t_1/2_: terminal half-life
^a^T_max_: time of C_max_
SD: standard deviation, ^a^Mean difference (90% CI) between the test and the comparator epoetin alfa
ANOVA: analysis of variance
ΔAUEC: area under the baseline-adjusted effect curve
ΔE_max_: maximum effect change
^b^T_max_: time of E_max_

## Supplementary information

Not applicable.

## Acknowledgements

The study was funded by Globe Biotech Limited. We thank Md. Harunur Rashid, the chairman of Globe Pharmaceuticals Group of Companies, Ahmed Hossain, Md. Mamunur Rashid, Md. Shahiduddin Alamgir and Abdullah Al Rashid, the directors of Globe Pharmaceuticals Group of Companies for their continuous support and encouragement. We also thank Md. Raihanul Hoque, Dibakor Paul, Biplob Biswas, Mithun Kumar Nag, Zahir Uddin Babor, GM Sajib Hassan, and Mijanur Rahman for their support for information and facility management system.

## Author contributions

Conceptualization: Kakon Nag and Naznin Sultana; Test product manufacturing and evaluation: Samir Kumar, Md. Enamul Haq Sarker, Md. Mashfiqur Rahman Chowdhury, Sourav Chakraborty, Md. Bipul Kumar Biswas, Md. Emrul Hasan Bappi, Ratan Roy, Maksudur Rahman Khan, Rony Roy and Mohammad Mohiuddin; methodology and supervision of clinical trial: Kakon Nag, Naznin Sultana and Mohammad Mohiuddin; execution of clinical trial: Mamun Al Mahtab, Sitesh Chandra Bachar, Md. Abdur Rahim and Md. Helal Uddin; manuscript writing and editing: Kakon Nag, Naznin Sultana, Mohammad Mohiuddin, Samir Kumar, Mamun Al Mahtab, Sitesh Chandra Bachar and Md. Abdur Rahim; project administration: Kakon Nag and Md. Helal Uddin; All authors have read and agreed to the manuscript.

## Availability of data and materials

The data that support the findings of this study are available within the article and its Supplementary document file, or are available from the corresponding author upon reasonable request.

## Declarations

### Ethics approval and consent to participate

The protocol for the study has got ethical clearance from the institutional review board (IRB; The Ethics Committee of Farabi General Hospital Ltd.) and approved by the Directorate General of Drug Administration (DGDA) of Bangladesh Ref. No. DGDA/CTP-1/06/2016/9916. The Clinical Research Organization (CRO Ltd.) conducted this investigation at Farabi General Hospital, Dhanmondi R/A, Dhaka 1209, in compliance with the principles of the Declaration of Helsinki and the International Conference on Harmonization’s Guideline for Good Clinical Practice.

The informed consent form (ICF), along with the protocol and methodology, was also approved by IRB and DGDA. Volunteers were given thorough information about the risks and benefits of the study and they have signed the ICF to affirm their willingness to voluntarily participate in the study. The trial protocol has been submitted and registered with the clinicaltrial.gov of the National Library of Medicine of the USA.

## Consent for publication

Not applicable.

## Author details

Not applicable.

